# Mosquito exposure and malaria morbidity; a micro-level analysis of household mosquito populations and malaria in a population-based longitudinal cohort in western Kenya

**DOI:** 10.1101/19008854

**Authors:** Wendy Prudhomme O’Meara, Ryan Simmons, Paige Bullins, Betsy Freedman, Lucy Abel, Judith Mangeni, Steve M. Taylor, Andrew A. Obala

**Affiliations:** Duke Global Health Institute, Duke University, Durham, NC 27708; Department of Medicine, School of Medicine, Duke University, Durham, NC 27708; School of Public Health, College of Health Sciences, Moi University, Eldoret, Kenya; Department of Biostatistics and Bioinformatics, School of Medicine, Duke University, Durham, NC 27708; Academic Model Providing Access to Healthcare, Moi Teaching and Referral Hospital, Eldoret, Kenya; School of Nursing, College of Health Sciences, Moi University, Eldoret, Kenya; School of Medicine, College of Health Sciences, Moi University, Eldoret, Kenya

**Keywords:** malaria, anopheles, cohort, insecticide treated net

## Abstract

**Background:** Malaria morbidity is highly overdispersed in the population. Fine-scale differences in mosquito exposure may partially explain this heterogeneity. However, exposure variability has not been related to individual malaria outcomes.

**Methods:** We established a cohort of 38 households to explore the effect of household-level mosquito exposure and individual insecticide treated net(ITN) use on relative risk(RR) of diagnostically-confirmed malaria. We conducted monthly active surveillance (n=254; 2,624 person-months) and weekly mosquito collection in all households (2,092 household-days of collection). We used molecular techniques to confirm human blood feeding and exposure to infectious mosquitoes.

**Results:** Of 1,494 female anopheles (89.8% *Anopheles gambiae s*.*l*.). 88.3% were fed, 51.9% had a human bloodmeal, and 9.2% were sporozoite-infected. 168 laboratory-confirmed malaria episodes were reported (incidence rate 0.064 episodes per person-month at risk, 95% confidence interval [CI]:0.055,0.074). Malaria risk was directly associated with exposure to sporozoite-infected mosquitoes (RR=1.24, 95%CI:1.11,1.38). No direct effect was measured between ITN use and malaria morbidity, however, ITN use did moderate the effect of mosquito exposure on morbidity.

**Conclusions:** Malaria risk increases linearly with vector density and feeding success for persons with low ITN use. In contrast, malaria risk among high ITN users is consistently low and insensitive to variation in mosquito exposure.

**Summary:** In this study, we measure the relationship between fine-scale spatio-temporal heterogeneity in exposure to infected and successfully-fed malaria vectors, the incidence of malaria, and their interaction with ITN use in a population-based cohort.

## Background

Cases of malaria are highly over-dispersed in human populations [1-4]: as little as 20% of individuals experience 80% of the disease burden. The reason for this heterogeneity is undoubtedly multifactorial, and even after adjusting for age and behavioral differences such as ITN use, the imbalance remains [2-4]. Quantifying the heterogeneity attributable to variability in entomological exposure requires fine-scale measurement of both mosquito populations and malaria incidence at a household or individual level. This has rarely been done and instead human cohort studies that incorporate exposure substitute either local infection prevalence in humans as a proxy for exposure [3, 5] or extrapolate entomological indicators from sentinel villages or households [4, 6-9]. Both approaches have limitations with regards to understanding the relationship between exposure and disease risk owing to the fact that disease risk is heterogeneous below the village level [10-13]. Such studies present inconsistent findings regarding exposure and incidence. Although cross-sectional studies of vector biting heterogeneity demonstrate significant over-dispersion in exposure to mosquito bites, this has not been linked to subsequent infection or disease distribution [14].

Recent studies support the role of mosquito exposure heterogeneity in understanding evolving malaria transmission dynamics, particularly in areas where endemicity is declining [14-18]. Mathematical models predict that exposure heterogeneity could reduce the effectiveness of malaria control measures [19]. Linking individual mosquito exposure to morbidity outcomes would enable more accurate estimation of the effects of vector control interventions at the population level. However, cohort studies rarely measure fine-scale entomological exposure.

Here we offer a high-resolution picture of individual-level incidence of symptomatic disease related to weekly household mosquito exposure metrics. We conducted a longitudinal study of 36 households in a moderate-transmission setting in Western Kenya in which we collected weekly indoor-resting mosquitoes and passively-detected cases of malaria. We hypothesized that excess variability in individual malaria risk would be at least partly explained by more precise measures of mosquito exposure and that the relationship between disease and mosquito exposure would increase with increasing specificity of exposure metric; from vector abundance, to feeding success, to human biting rate, and finally vector infectiousness.

## Methods

### Study site

The study was carried out in Webuye East and West sub-counties located in western Kenya approximately 400 kilometers northwest of Nairobi and 60 from the border with Uganda. The sub-counties are rural and most families engage in small-scale farming and animal husbandry. Very few families have access to amenities such as municipal water or electricity (<5%) and 53% live below the poverty line [20]. There is one small town center along the main highway that bisects the county. Malaria transmission is moderate and perennial with two seasonal peaks after the long rains (May-June) and the short rains (Sept-October) although the timing and intensity of the transmission peaks can vary from year to year. Vector control interventions are limited to ITN distribution and use; IRS has not taken place in this area and the use of individual protective measures such as sprays and coils is very uncommon.

### Cohort

In July 2017, we established a cohort of 36 households in three villages that exhibited different levels of malaria transmission based on previous work [21, 22]. We enrolled an index household at random and then neighboring households radiating outwards until 12 households were enrolled per village in a natural grouping. Two households were replaced with neighboring households when the entire household migrated. All residents older than 12 months who regularly slept in the household were enrolled. Children <12 months at baseline were enrolled after they passed their first birthday. A household is defined as a group of people who share cooking arrangements and may consist of one or more buildings grouped together in a compound. Information about building construction, sleeping spaces, and ITN ownership was collected at enrollment.

### Cohort surveillance

At baseline, we registered all household members and collected dried blood spots from finger pricks. Each month, we recorded data about bednet use in the preceding week and illness in the preceding one month from all consenting household members over the age of 12 months. In addition, all household members were invited to contact the study team whenever they experienced a malaria-like illness. When contacted, study staff visited the home to test the participant for malaria using a rapid diagnostic test (Carestart © Malaria HRP2 *Pf* from Accessbio). If the participant had a negative test, they were referred with the test results to the nearest health facility. If the test was a positive, the participant collected quality-assured ACT from the local pharmacy at no charge to the participant.

### Mosquito collection and identification

Once per week, on the same day each week, the study team visited each household early in the morning to collect indoor resting mosquitoes via aspiration with Prokopacks (John W. Hock Company). Participants were asked to leave windows and doors closed until the team arrived. Mosquitoes were stored in collection cups in coolboxes with icepacks until they were transported back to the laboratory. Mosquitoes were separated by genus and sex, and females were graded according to their abdominal status. Female Anopheles were imaged under magnification for post-hoc speciation, first by study staff and then by a senior entomologist in a subset of 25%. After imaging, mosquitoes were dissected between the thorax and abdomen, and these parts were stored in separate tubes packed with desiccant at room temperature.

### Molecular detection of parasites and human blood in mosquitoes

Mosquito specimens were manually disrupted in individual tubes and then genomic DNA was extracted using a Chelex-100 protocol. Each gDNA extract was tested in a duplex TaqMan real-time PCR assay targeting both *P. falciparum* and human beta-tubulin [23]. Samples were tested in duplicate in 384-well plates on an ABI Quantstudio 6 platform; threshold lines were set manually in the exponential phase of amplification. Reaction plates were prepared in dedicated workspaces with filtered pipet tips, and each included *P. falciparum* gDNA and human DNA as positive controls and molecular-grade water as a negative control. Genomic DNA was extracted from human dried blood spots and tested in the same real-time PCR assay.

### Data analysis

The primary outcome was malaria, which was defined as self-reported illness leading to a positive malaria RDT result in the preceding 30 days. Test results were confirmed by reviewing the medical record whenever possible. Following a positive test, individuals were censored for 21 days. The primary objective of our analysis was to estimate the impact of household-level mosquito indices on an individual’s risk of malaria. We investigated the impact of four different mosquito indices, calculated for each household in each month as the sum of the counts from weekly mosquito collection visits: the total number of 1) female Anopheles, 2) blood-fed female Anopheles, 3) female Anopheles mosquitoes collected with human blood detected by PCR in the abdomen, and 4) female Anopheles with P falciparum sporozoites detected in the head. Due to collinearity between the exposures (e.g. the number of sporozoite positive mosquitoes being a subset of the total number of mosquitoes collected, etc.), separate models were fit for the effect of each exposure on each morbidity outcome.

We used generalized linear mixed models (GLMM), with the outcome defined as the presence or absence of malaria for each individual at each follow-up month. To estimate relative risks, we used a robust (modified) Poisson model [24]. To account for clustering by household, the model included a random intercept on household with a compound symmetric covariance structure (note that this assumes household clustering is constant over time). To account for clustering within individual over time, the model included a random intercept on individual, nested within household, with a homogenous first-order autoregressive covariance structure. Each model included a fixed-effect for village, age at baseline (coded as a categorical variable with five-levels: <=1, 1-5, 6-10, 11-17, and >=18 years), gender, and number of nights of ITN use in the past week (dichotomized as >=6 and <6 nights). Additionally, we investigated interactions between the mosquito exposure variables and ITN use.

In order to quantify the amount of variability in individual incidence that could be explained by the model covariates, we sequentially estimated marginal R^2^ for generalized linear mixed models, following the method of [25]. Marginal R^2^ allows for assessment of the impact of the fixed effects independent of the random effect structure of the model. Starting with the null model, we added covariates sequentially in order of the magnitude of their fixed effect from the full model, until all demographic adjustment factors were included. Finally, each mosquito exposure variable, and its interaction with ITN use, were added one at a time to this adjusted model.

Secondary exploratory objectives included investigating the impact of including in the model covariates related to individual travel history (e.g. using a bed-net while traveling, traveling to a malaria endemic area, etc.), the number of hours spent under a bed-net each night, and the number of individuals living in the household.

### Research ethics and consent

All heads of household were asked to consent for participation in mosquito collection and household data collection. Each individual provided consent for monthly dried blood spot collection and RDT testing. A parent or legal guardian provided consent on behalf of a child under 18 years of age. Children ten years and above were also asked to provide assent for participation. The study was reviewed and approved by Duke University Institutional Review Board and Moi University and Moi Teaching and Referral Hospital Institutional Research Ethics Committee.

## Results

From June 12, 2017 until July 31, 2018, 254 participants from 38 households were followed by monthly active surveillance and weekly entomological surveillance for a total of 2,624 person months of observation. The cohort demographics are described in Table 1. Sixty percent of participants were children less than 18 years of age at enrollment and each person provided an average of 11.1 months of follow-up. At enrollment, 54.9% (133/242) of participants were infected with *P. falciparum* as detected by a highly-sensitive PCR assay. 72% of participants had an ITN for their sleeping space; this proportion was slightly lower in Village S (59%) compared to Village K and M (82% and 78%, respectively). In 76.7% (2,013/2,624) of monthly follow-ups, participants reported sleeping under their net each day of the previous seven days.

**Table 1:** Participant characteristics, baseline bednet use, follow-up time and malaria episodes reported by village and total numbers

|  | Village K | Village M | Village S | Total |
| --- | --- | --- | --- | --- |
| Demographics |  |  |  |  |
| Households | 12 | 13 | 13 | 38 |
| Participants | 80 | 78 | 96 | 254 |
| Median Household size<br>(range) | 6.5 (5-13) | 6 (3-15) | 8 (3-14) | 6.5 (3-15) |
| Female | 45 (51.7%) | 46 (57.5%) | 57 (57.0%) | 148 (55.4%) |
| <1 years | 2 (2.5%) | 3 (3.9%) | 5 (5.2%) | 10 (3.9%) |
| 1-5 years | 13 (16.3%) | 12 (15.4%) | 9 (9.4%) | 34 (13.4%) |
| 6-10 years | 17 (21.3%) | 16 (20.5%) | 24 (25.0%) | 57 (22.4%) |
| 11-17 years | 16 (20.0%) | 13 (16.7%) | 22 (22.9%) | 51 (20.8%) |
| 18 and above | 32 (40.0%) | 34 (43.6%) | 36 (37.5%) | 102 (40.2%) |
| ITN coverage |  |  |  |  |
| Sleeps in space with an ITN<sup>a</sup> | 64/78 (82%) | 59/76 (78%) | 54/92 (59%) | 177/246 (72%) |
| Sleeps in space with untreated net<sup>a</sup> | 3/78 (3.8%) | 0 | 0 | 3/246 (1.2%) |
| Slept under ITN last night<br>(baseline) | 67/80<br>(83.8%) | 58/78<br>(74.4%) | 60/96<br>(62.5%) | 185/254<br>(72.8%) |
| Observations |  |  |  |  |
| Person months | 879 | 808 | 946 | 2633 |
| Average person months per<br>participant (SD) | 11.42 (3.69) | 11.22 (7.60) | 10.63 (4.08) | 11.06 (5.29) |
| Malaria morbidity |  |  |  |  |
| Suspected malaria illness<br>events | 136 | 145 | 152 | 433 |
| Suspected malaria episodes<br>with positive malaria test | 51 (37.5%) | 55 (37.9%) | 62 (40.8%) | 168 (38.8%) |

<sup>a</sup>2 people in Village K, 2 in village M and 4 in village S were missing sleeping space information

### Mosquito indices

Across 2,092 household-days, we collected a total of 1,494 female anopheles mosquitoes (Table 2, Figure 1). Nearly 90% (n=1,023) were identified as *Anopheles gambiae* group and this proportion varied only slightly across the villages. Village M recorded the highest number of malaria vectors overall (66.5 per household) and the highest proportion of *An. funestus* (9.3%). Other Anopheles spp. were observed at low frequencies. By microscopic evaluation, the majority of mosquitoes had recently fed: 11.2% (n=168) were freshly bloodfed, 31.9% (n=477) were half gravid, and 39% (n=587) were gravid. In PCR testing, we detected human blood in 51.9% of all abdomens (n=776), including 79.8% (n=134) of the freshly bloodfed specimens. The prevalence of *P. falciparum* sporozoites in the heads of female Anopheles was 9.2% (n=134). The proportions of mosquitoes with human blood or sporozoites did not differ by species or village.

**Figure 1:**
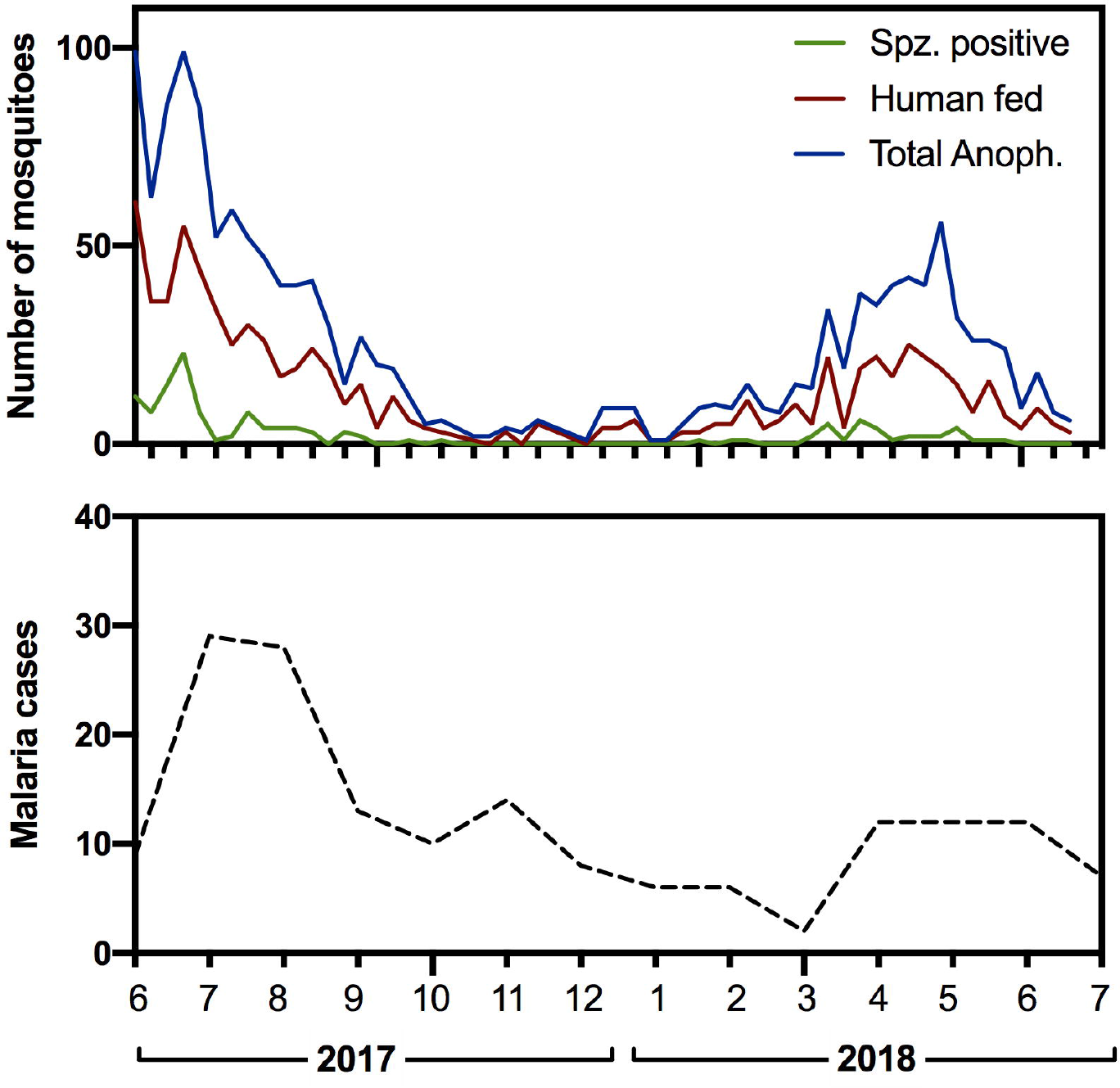
Weekly entomological exposures (a) and monthly confirmed malaria cases (b) from June 2017 to July 2018

**Table 2:** Abundance and characteristics of anopheles mosquitoes collected in the cohort households

|  | Village K | Village M | Village S | Total |
| --- | --- | --- | --- | --- |
| No. of households |  |  |  |  |
| Total female *Anoph.* collected | 404 | 858 | 232 | 1494 |
| Anopheles per HH, mean (range) | 33.7 (8-72) | 66 (7-198) | 17.8 (4-45) | 38.4 (4-201) |
| Microscopic analysis, no. (%) |  |  |  |  |
| Unfed | 67 (16.7%) | 81 (9.4%) | 20 (8.6%) | 168 (11.2%) |
| Freshly fed | 111 (27.6%) | 297 (34.6%) | 69 (29.7%) | 477 (31.9%) |
| Half gravid | 165 (41.0%) | 320 (37.3%) | 102 (44.0%) | 587 (39.3%) |
| Gravid | 58 (14.4%) | 158 (18.4%) | 39 (16.8%) | 255 (17.1%) |
| Undetermined | 3 (0.7%) | 2 (0.2%) | 2 (0.9%) | 7 (0.5%) |
| Human blood in abdomen by PCR, % (n/N) | 53.1<br>(207/390) | 53.5<br>(447/835) | 54.7<br>(122/223) | 53.6<br>(776/1448) |
| *P. falciparum* sporozoites, % (n/N) | 10.2<br>(40/393) | 9.3<br>(78/839) | 7.0<br>(16/230) | 9.2<br>(134/1462) |
| Species<sup>1</sup> | 319 | 631 | 189 | 1139 |
| *An. gambiae* s.l | 298 (93.4%) | 544 (86.2%) | 181 (95.8%) | 1023 (89.8%) |
| *An. funestus* complex | 14 (4.4%) | 55 (8.7%) | 2 (1.1%) | 71 (6.2%) |
| *An. demeilloni* | 3 (0.9%) | 6 (1.0%) | 3 (1.6%) | 12 (1.1%) |
| *An. pretoriensis* | 1 (0.3%) | 11 (1.7%) | 1 (0.5%) | 13 (1.2%) |
| Other<sup>2</sup> | 3 (1.0%) | 15 (2.4) | 2 (1.1%) | 20 (1.8%) |

<sup>1</sup> 355 mosquitoes were not identified to species due to technical problems with the microscope.
Totals and percentages here only include identified mosquitoes.
<sup>2</sup> Other species include *An. maculipalpis* (3), *An. salbaii* (4), *An. rufipes* (2), *An. dancalicus* (5), *An. coustani* (6)

### Malaria incidence

During 2,624 person-months of follow-up, there were 168 cases of malaria in 107 people (Figure 1), yielding an overall incidence rate of 0.064 (95% CI: 0.055, 0.075) cases per person-month. Among these 107 people, 66 (61.7%) reported only a single episode, 26 (24.3%) reported two episodes, and 15 (14.0%) reported 3 or more episodes (the maximum was 5). The median number of days between sequential episodes was 37 (IQR: 62). Overall, 30% (n=74) of participants experienced 80% (n=135) of episodes (Supplemental Figure 1) and 58% (n=147) of participants never experienced an episode of malaria.

### Associations between mosquito indices and malaria

We estimated the relative risk of malaria in models without and with each of our mosquito indices (Table 3). As expected, the risk of malaria declined with increasing age, and there was no association with gender (RR 1.00; 95% CI 0.72 – 1.40). Relative to high ITN users, those with low use were at similar risk (RR 1.16; 95% CI 0.76 – 1.79). In models with individual mosquito indices, malaria risk was not significantly associated with the monthly household abundance of malaria vectors (RR 1.02; 95% CI 0.97 – 1.08) nor the number of mosquitoes who had human blood in their abdomen (RR 1.04; 95% CI 0.97 – 1.12). However, the risk of malaria increased by 24% with each additional infectious mosquito collected the previous month (RR 1.24; 95% CI 1.11 – 1.38).

**Table 3:** Relative risk of malaria in the prior 30d in base model and in models including mosquito indices. The effect of monthly individual ITN use and monthly household-level mosquito exposure on the relative risk of malaria in the last 30 days. Each model is adjusted for household and individual-level demographic characteristics.

|  | Base model | Individual mosquito index models |  |  |  |
| --- | --- | --- | --- | --- | --- |
|  |  | 1 | 2 | 3 | 4 |
| Village<br>(Kinesamo vs. Sitabich) | 0.70<br>(0.43,1.14) | 0.68<br>(0.42,1.10) | 0.68<br>(0.42,1.11) | 0.68<br>(0.42,1.1) | 0.63<br>(0.39,1.00) |
| Village<br>(Maruti vs. Sitabich) | 0.91<br>(0.51,1.62) | 0.81<br>(0.43,1.53) | 0.81<br>(0.43,1.55) | 0.81<br>(0.43,1.51) | 0.77<br>(0.41,1.42) |
| <1 yrs | 1.63<br>(0.45, 5.86) | 1.63<br>(0.45, 5.82) | 1.62<br>(0.45, 5.81) | 1.62<br>(0.45, 5.82) | 1.60<br>(0.45, 5.72) |
| 1-5 yrs | **2.10**<br>**(1.29, 3.40)** | **2.08**<br>**(1.28, 3.38)** | **2.08**<br>**(1.28, 3.38)** | **2.08**<br>**(1.28, 3.38)** | **2.09**<br>**(1.29, 3.38)** |
| 6-10 yrs | 1.55<br>(0.99, 2.42) | **1.58**<br>**(1.01, 2.47)** | **1.58**<br>**(1.01, 2.47)** | **1.58**<br>**(1.01, 2.48)** | **1.60**<br>**(1.03, 2.49)** |
| 11-17 yrs | 1.16<br>(0.66, 2.02) | 1.17<br>(0.67, 2.03) | 1.17<br>(0.67, 2.03) | 1.17<br>(0.67, 2.03) | 1.19<br>(0.68, 2.07) |
| 18+ yrs | *Ref* | *Ref* | *Ref* | *Ref* | *Ref* |
| Male (vs. Female) | 1.00<br>(0.72,1.40) | 1.01<br>(0.72,1.42) | 1.01<br>(0.72,1.42) | 1.01<br>(0.72,1.42) | 1<br>(0.72,1.39) |
| High ITN use (vs. Low) | 1.16<br>(0.76,1.79) | 1.21<br>(0.78,1.87) | 1.21<br>(0.78,1.88) | 1.21<br>(0.79,1.86) | 1.43<br>(0.91,2.24) |
| No. of household members | **0.91**<br>**(0.83,0.99)** | **0.91**<br>**(0.83,0.99)** | **0.91**<br>**(0.83,0.99)** | **0.91**<br>**(0.83,0.99)** | **0.9**<br>**(0.82,0.98)** |
| **No. of Anoph. mosquitoes** |  | 1.02<br>(0.97,1.08) |  |  |  |
| **No. Bloodfed Anoph. mosquitoes** |  |  | 1.03<br>(0.97,1.08) |  |  |
| **No. Human bloodfed Anoph. mosquitoes** |  |  |  | 1.04<br>(0.97,1.12) |  |
| **No. Sporozoite positive mosquitoes** |  |  |  |  | **1.24**<br>**(1.11,1.38)** |

Because high ITN use was not associated with a reduced risk of malaria, we hypothesized that ITN use may modify the effect of mosquito exposure on malaria risk. We tested this hypothesis by examining the interaction between ITN use and each mosquito exposure variable (Table 4). We demonstrate significant correlation between both mosquito abundance and malaria risk, and mosquito feeding success and malaria risk, in models that include the exposure:ITN interaction term. The relative risk of malaria among high ITN users compared to low ITN users decreased by 6% (RR=0.94, 95%CI: 0.89-0.99) for each additional anopheles mosquito in the household and by 10% (RR=0.90, 95%CI: 0.82-0.99) for each additional human-fed mosquito. ITN use now appears positively correlated with malaria risk in models that include the interaction term, however, we can understand this when we examine the results graphically (Figure 2). At low mosquito abundance, individuals with high ITN use have equal or slightly elevated malaria risk compared to those who have lower ITN use. However, the risk of malaria increases linearly with increasing mosquito exposure among those with low ITN use, but remains low and constant for those who use an ITN consistently. Similar trends are seen for mosquito feeding success and human feeding success.

**Figure 2:**
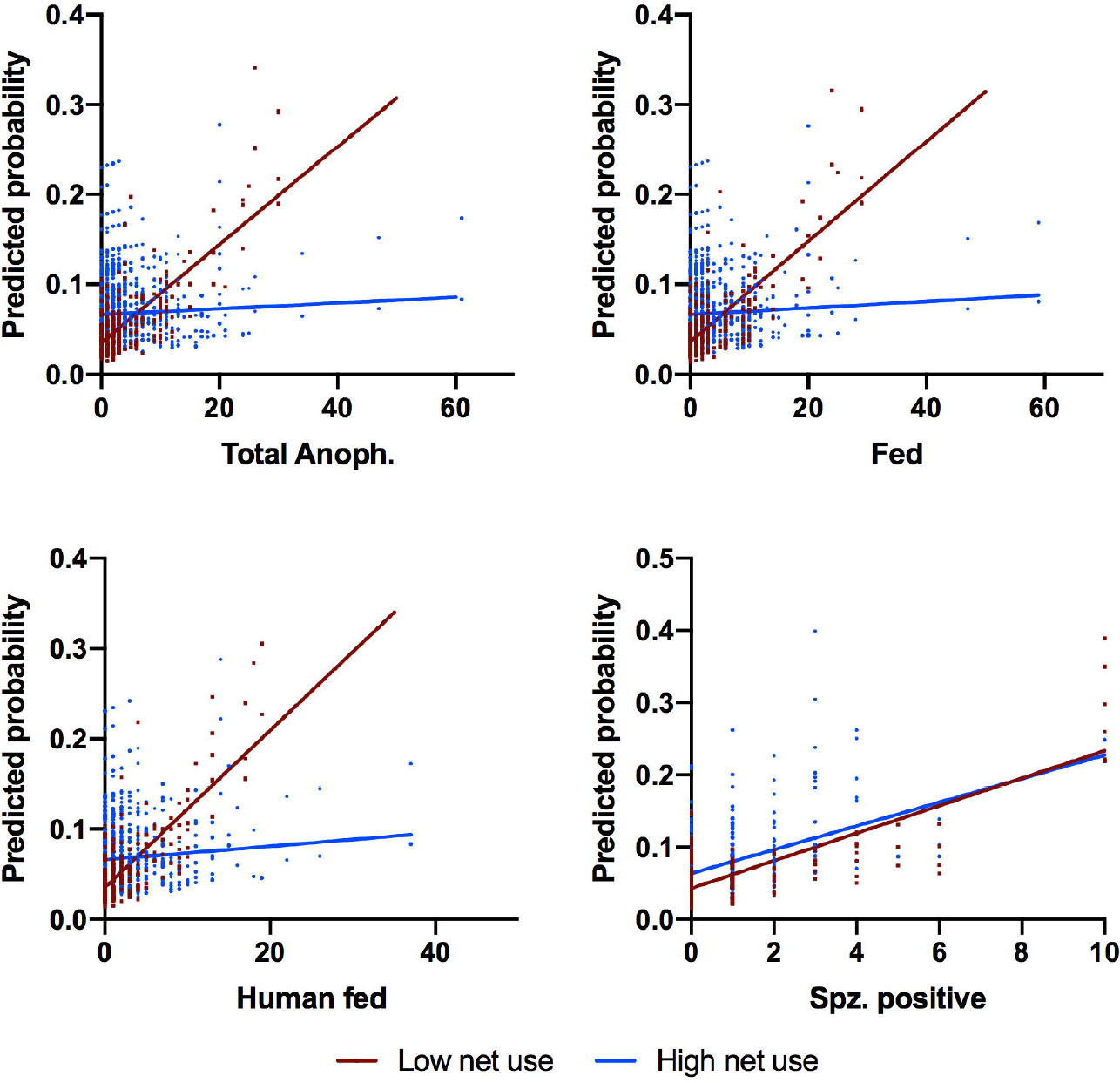
Predicted probability of malaria episode by mosquito exposure for high versus low net use

**Table 4:** The effect of individual ITN use, mosquito exposure, and the interaction between ITN use and mosquito exposure on the relative risk of malaria in the last 30 days. Each column represents a separate model adjusted for age, household size, gender, and village.

| Relative risk of malaria<br>in the last 30 days | Model 1 | Model 2 | Model 3 | Model 4 |
| --- | --- | --- | --- | --- |
| ITN Use<br>(High vs. Low) | 1.69<br>(1.05, 2.72) | 1.66<br>(1.04, 2.64) | 1.71<br>(1.07, 2.74) | 1.44<br>(0.90, 2.30) |
| Total # of Anoph.<br>Mosquitoes | 1.08<br>(1.013 1.12) |  |  |  |
| # of Fed Anoph.<br>Mosquitoes |  | 1.08<br>(1.03, 1.13) |  |  |
| # of Anoph. Mosquitoes<br>Fed on Human Blood |  |  | 1.13<br>(1.04, 1.22) |  |
| # of Sporozoite Positive<br>Anoph. Mosquitoes |  |  |  | 1.24<br>(1.14, 1.36) |
| ITN Use * Exposure<br>Interaction | 0.94<br>(0.89, 0.99) | 0.94<br>(0.88, 0.99) | 0.90<br>(0.82, 0.99) | 0.99<br>(0.85, 1.16) |

In analyses of marginal R^2^ with incorporation of mosquito indices in malaria risk models, age, household size, and exposure to sporozoite-infected mosquitoes accounted for the largest increase in R^2^ (2.6%, 2.9% and 2.3%, respectively; Figure 3). Other mosquito exposures accounted for only a very small proportion of the heterogeneity. Despite the small absolute change in R^2^, it is interesting to note that household exposure to infectious mosquitoes and age contribute nearly equally to explaining the distribution of malaria incidence in the population.

**Figure 3:**
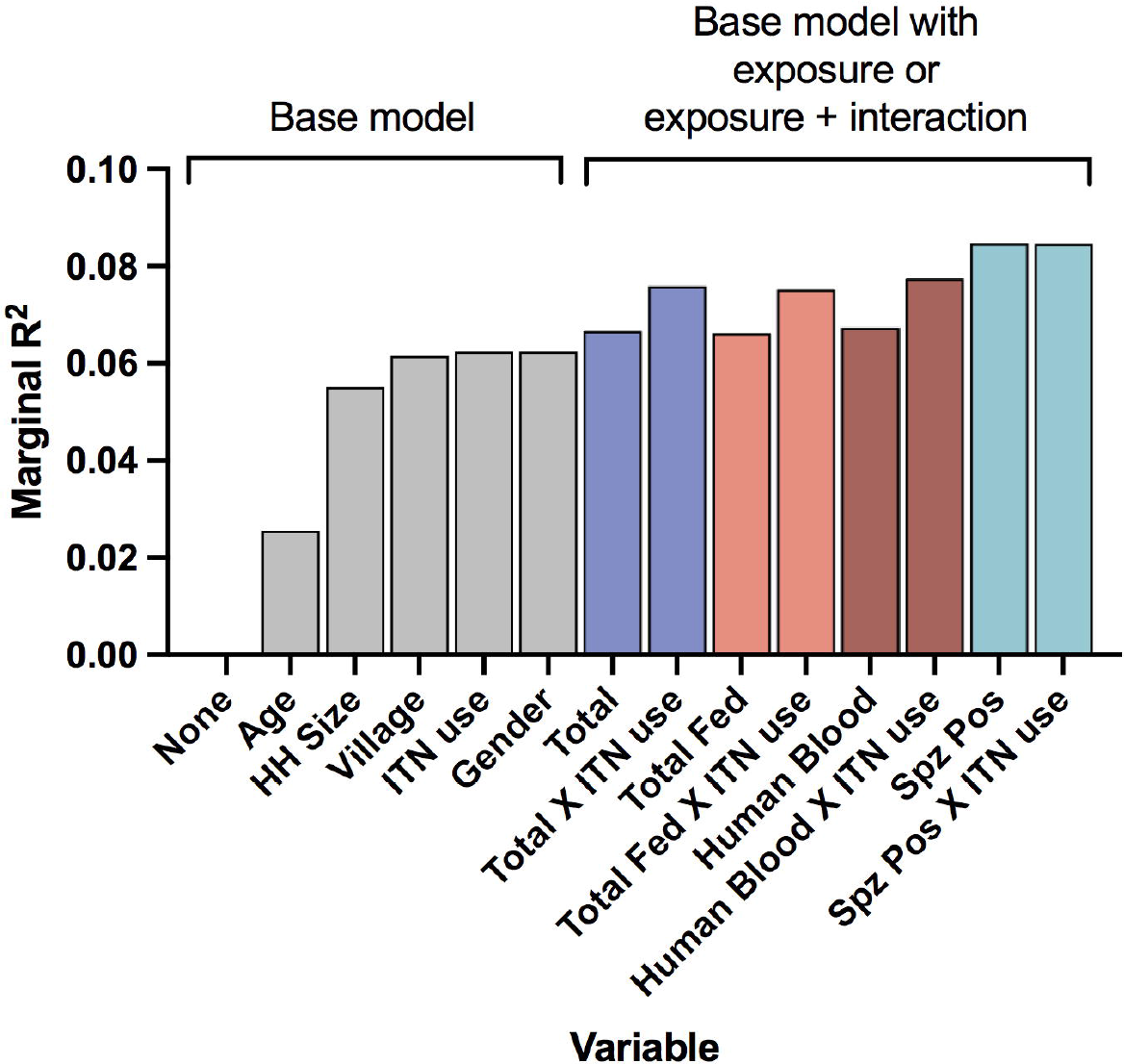
Estimated marginal R^2^ for models of malaria incidence where covariates are added sequentially (base model) to measure the effect on model fit. Separate models are fit with base model plus one exposure variable or one exposure variable with interaction on net use.

## Discussion

In this high-resolution paired entomological and epidemiological cohort, we measured weekly household-level mosquito exposure alongside laboratory-confirmed malaria episodes in a population-based sample of 254 individuals in 38 households. We show that, among putative entomological indices, the risk of malaria in humans is mediated by neither the abundance of mosquitoes overall nor those that are blood-fed mosquitoes but rather only by the risk of malaria in mosquitoes (sporozoites). We further demonstrate an interaction between the effect of ITN use and mosquito exposure on malaria episodes, in that, compared to suboptimal ITN use, high ITN use attenuates the deleterious effects of vector density and feeding success on the risk of malaria.

The relationship between mosquito exposure and human malaria risk is difficult to quantify due to the time lag between an infectious bite and patent infection, and the uncertainty in assigning individual exposure based on entomological variables. Some studies compare population based measures of disease prevalence or incidence with vector density observed at sentinel locations and are essentially ecological comparisons of two populations over time [6, 15]. Alternatively, values from sentinel households are locally extrapolated to unmeasured households [4, 7-9]. Although we fall short of assigning individual exposure, which may vary within a household [26, 27], our weekly household-level entomological measurements give new insight into the nature of the relationship between mosquito exposure and disease and a means to estimate the change in risk under different exposure control strategies. For example, since both the effect of mosquito abundance and feeding success on malaria morbidity are moderated by individual ITN use, we can estimate a personal protective efficacy of ITN use of up to 30% as exposure increases. In households where as few as 8 mosquitoes were collected in a month, ITNs offer measurable protection relative to those who do not use an ITN. The low, but non-zero, malaria risk experienced by consistent ITN users may reflect exposure outside of the net, due to modified mosquito behaviors such as biting early in the evening or outdoors [28-32].

Our findings relating risk of malaria to basic demographic factors are consistent with previous studies. We and others [3, 4, 33-35] measure declining incidence of clinical malaria with increasing age. The observation that increasing household size is associated with decreasing individual risk can be interpreted as reduced individual risk of exposure when more hosts are available. Although individual ITN use has been associated with reduced risk of malaria in longitudinal studies of young children in the context of high transmission and low ITN coverage [1, 13], we and others fail to detect a direct, individual protective effect [3, 7]. Overall, behavioral and demographic differences explain relatively little of the heterogeneity in malaria cases. After incorporating entomological exposures and the effect of ITN use on those exposures, the proportional amount of heterogeneity explained increased by 36%, however the absolute value of R^2^ remained small.

There are notable limitations to this study. First, the time span is relatively short (13 months); a longer duration of observation would be desirable to capture important annual variation in transmission and exposure. Second, the malaria morbidity outcome is based on recall over a one-month period. However, we mitigated possible recall bias and missing information by reviewing medical records to confirm test results and ensuring affordably, timely and accurate malaria diagnosis was available to participants.

Despite these limitations, we demonstrate that infections in mosquitoes within a household are a direct risk factor for infections in individuals. This risk is not mitigated by ITN use. Other entomological exposures measured at the household-level, including malaria vector density and human-fed anopheles, increase the risk of malaria episodes differentially in individuals who do not use an ITN consistently. We show that individual protective efficacy of ITNs scales with exposure and fine-scaled entomological measures can explain as much heterogeneity in malaria risk in the population as age.

## Data Availability

Data will be available upon final publication of the peer-reviewed version of the manuscript

## Acknowledgements

We would like to thank our exceptional field team (I. Khaoya, L. Marango, E. Mukeli, E. Nalianya, J. Namae, L. Nukewa, E. Wamalwa, and A. Wekesa) and all the families who gave their valuable time for this study.

